# Multi-Omics Analysis of Red Blood Cells Reveals Molecular Pathways Underlying Thalassemia Severity Beyond Globin Gene Mutations

**DOI:** 10.1101/2025.02.09.25321939

**Authors:** Nibedita Mitra, Upasana Bhattacharyya, Prosanto Chowdhury, Arijit Pal, Arvind Korwar, Samsidhhi Bhattacharjee, Anupam Basu

## Abstract

**Background:** Hemoglobinopathies are the most common monogenic genetic disorders, primarily managed through blood transfusions or bone marrow transplantation. Clinical severity other than mutational effect not well investigated and still unknown. This study aims to identify dysregulated molecular pathways in red blood cells contributing to thalassemia severity.

**Method:** From a cohort of 285 hemoglobinopathy patients, 10 age-matched individuals with identical compound heterozygous mutations (IVS 1-5 G>C and CD 26 G>A) were screened. Five had severe thalassemia requiring regular transfusions, while five had a non-severe form requiring fewer transfusions. RNA sequencing and proteome analysis were conducted on isolated RBCs, through Novaseq and Orbitrap MS platform respectively. Bioconductor-R and different bioinformatics tools were utilized subsequently.

**Results:** Transcriptome analysis revealed an increased percentage of snRNA transcripts in all over thalassemia group. Pathways related to autophagy, mitophagy, and chaperone-mediated folding were enriched in the severe group. Thus, dysregulated genes, linked to ineffective erythropoiesis were fished out also.

**Conclusion:** In this this study, thalassemia subjects were of same mutational genotype, but clinically opposite severity. Accordingly, first time identified six pathways which are responsible for thalassemia severity independent of mutational burden. These dysregulated pathways can be further be explored and targeted experimentally for drug development.

## INTRODUCTION

Hemoglobinopathy, one of the most prevalent genetic disorders worldwide, arises from mutations, primarily in the beta-globin gene (*HBB*), leading to disrupted globin chain synthesis. This condition significantly impacts global child health, resulting in high mortality rates^1^. Although it is a monogenic Mendelian trait, its diverse phenotypic presentations pose challenges for effective treatment and management.

Thalassemia, a major subset of hemoglobinopathies, is categorized into two clinical groups based on disease severity: Transfusion-Dependent Thalassemia (TDT), or severe thalassemia, and Non-Transfusion-Dependent Thalassemia (NTDT), or non-severe thalassemia. Disease severity is largely determined by the type and burden of beta- or alpha-globin gene mutations. However, patients with identical *HBB* genotypes often exhibit varying clinical severities, indicating that factors beyond the primary genetic mutation, such as differential molecular pathways, contribute to this heterogeneity. Common symptoms of thalassemia include bone marrow expansion, extramedullary hematopoiesis, and hepatosplenomegaly, which worsen the disease burden ^2–5^.

Currently, regular blood transfusions remain the cornerstone of thalassemia management due to the lack of effective, targeted drugs. However, transfusions are associated with severe complications, including iron overload, cardiotoxicity, liver fibrosis, and increased risk of infections. Hydroxyurea, originally developed as an HbF inducer for sickle cell anemia, has been repurposed for thalassemia but shows limited efficacy, particularly in severe cases, and its success depends on specific genetic polymorphisms like Xmn1^6^. The recent approval of Luspatercept for thalassemia treatment showed limited efficacy^7–8^.

One of the key challenges in thalassemia drug development is identifying suitable molecular targets that can be leveraged for therapeutic intervention. Red blood cells (RBCs), essential for hemoglobin production, are profoundly affected in thalassemia due to ineffective erythropoiesis and premature destruction. Recent studies have identified RNA transcripts in normal human erythrocytes, shedding light on RBC biology^9–15^. However, the transcriptional and translational profiles of RBCs in thalassemia patients, particularly concerning disease severity, remain largely unexplored. Understanding these molecular alterations could provide critical insights for drug discovery. However, the understanding the molecular mechanisms of disease severity beyond the mutational burden of the globin gene is the key for further step for drug development. To address this, we analyzed RBCs directly from patients with divergent clinical severities but identical *HBB* genotypes. Thus, the present study aims to identify dysregulated pathways in the RBCs of severely affected thalassemia patients by comparing them with a non-severe cohort through transcriptional profiling and validating findings at the translational level. The goal is to uncover the molecular pathways that can be exploited for future study.

## RESULTS

### Cohort Presentation

In a cohort of hemoglobinopathy patients with varying genotypes, we focused specifically on those with the β+/β0 compound heterozygous genotype. Unlike other genotypes such as β0/β0 or β+/β+, the β+/β0 genotype presents with a phenotypic dichotomy, showing both severe and non-severe clinical manifestations. We excluded genetic variants of known secondary modifiers, such as mutations in the alpha-globin gene or the HbF-inducing Xmn1/γG-globin-185 C>T polymorphism. This ensured a homogeneous genotype among the subjects. From this group, patients with identical HBB genotypes—IVS 1-5 G>C and CD 26 G>A—matched for age and sex, were selected for multi-omics investigation (Table 1). Consequently, the study focused on ten thalassemia patients with identical mutational profiles but distinct phenotypic differences. Additionally, five age-matched healthy individuals were included as controls. According to the TIF guidelines ^16^, the ten thalassemia subjects selected were categorized into equal two groups: severe (TDT) and non-severe (NTDT), despite having the same genetic burden in terms of globin gene mutations. In the severe group, five patients presented very early in life (<12 months) with low hemoglobin levels of 6–7.5 g/dL, requiring regular blood transfusions at approximately two-month intervals. These patients underwent biochemical screening for iron overload (serum ferritin) and were placed on regular iron chelation therapy. The remaining five patients comprised the non-severe group, showing signs of anemia at a much later age and requiring fewer transfusions, if any, while maintaining an average hemoglobin level of 7–8 g/dL. As the serum ferritin levels indicated that most had iron overload within tolerable limits, they were not on regular iron chelation therapy

### Differential Gene Expression

#### Transcriptome Profile

Principal component analysis (PCA) and cluster heatmap analysis of the red blood cell (RBC) transcriptome revealed significant heterogeneity between thalassemia patients and healthy controls. A total of 2, 612 differentially expressed genes (DEGs) were identified when comparing thalassemia patients to controls. As per volcano plot, Among these, HSP90AB1, DEPP1, DYNLL1, HBG1, CFAP61, HBG2, HSPH1, FKBP4, MLLT3, and RBS12 emerged as the most dysregulated genes in the RBCs of thalassemia patients. The selection criteria for identifying DEGs were a log2 fold change (log2Fc) greater than 2 or less than -2 and an adjusted p-value (FDR) below 0.05 (Fig. 1A-C, Supplementary Table S1). To, gain insights into the biological implications of these DEGs in thalassemia, functional enrichment analyses were performed. Gene Ontology (GO) molecular function analysis revealed that the upregulated genes were significantly enriched in activities such as hemoglobin alpha binding, hemoglobin binding, haptoglobin binding, oxygen carrier activity, oxidoreductase activity, peroxidase activity, unfolded protein binding, and chaperone binding (Fig. 2A).Further analysis using Gene Set Enrichment Analysis (GSEA) demonstrated that the most positively enriched gene set was associated with ATP-dependent protein folding chaperones, with a Normalized Enrichment Score (NES) of 3.01, indicating a strong functional association (Fig. 2B). KEGG pathway enrichment analysis revealed a notable association of the upregulated DEGs with the ferroptosis pathway, suggesting a role for this cell death process in the pathophysiology of thalassemia (Supplementary File S1: Fig. S1B). Conversely, the downregulated genes in RBCs from thalassemia patients were enriched in functions related to IgG binding, interleukin-1 receptor binding, pattern recognition receptor binding, phosphotyrosine residue binding, phosphoprotein binding, actin filament binding, and actin binding. These findings indicate a potential disruption in immune and cytoskeletal functions in the RBCs of thalassemia patients.

**Figure 1.**
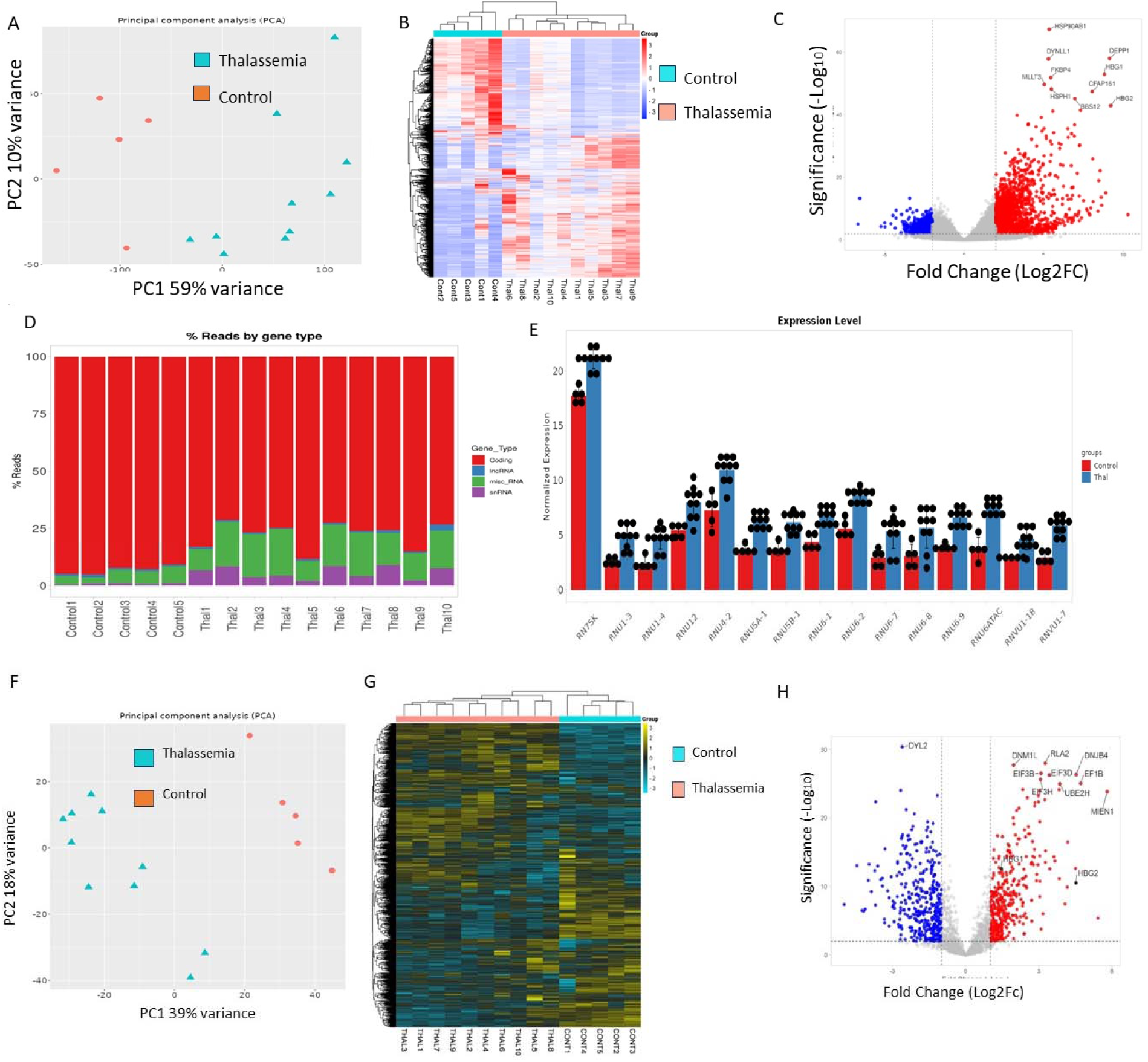
Analysis of RBC transcriptome and proteome data: A) Principal Component Analysis showing variance between patient and control group of RBC transcriptome data of ten Thalassemia patients and five control human subjects. B) Heatmap showing differential expression of top 2000 genes between patient and control groups. (C) Volcano plot showing most upregulated and down regulated genes in Thalassemia patient compared to control subjects (D) Bar graph representing the % of Reads by transcript type. E) Bargraph showing normalized expression levels for significant snRNAs were highly expressed in thalassemia group compared to control group. F) Principal Component Analysis showing variance between patient and control group of proteome data, G) Heatmap showing the differential abundance between patient and control groups. (H) Volcano plot showing most high and low abundant proteins in Thalassemia compared to control subjects.

**Figure 2.**
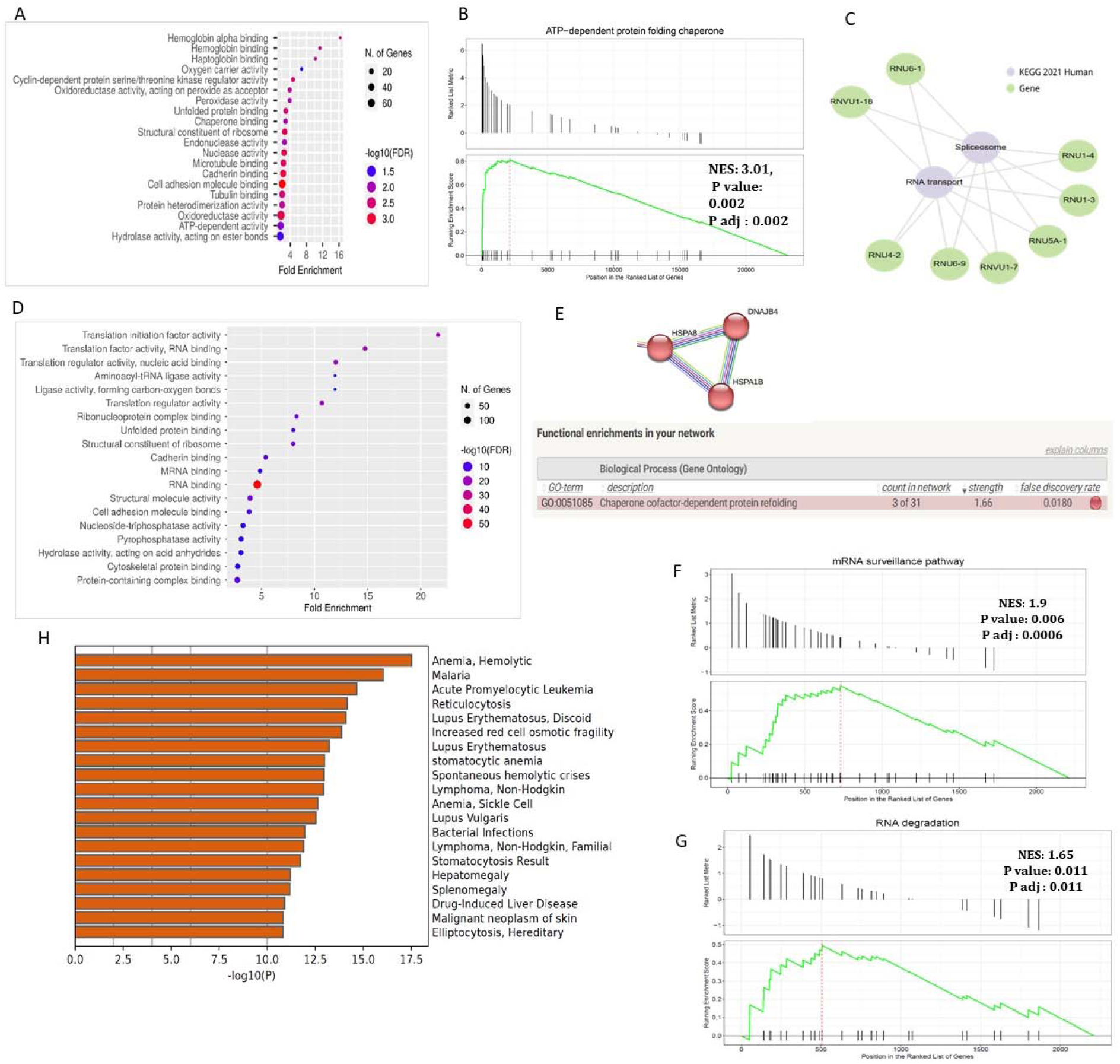
Functional Enrichment Analysis of Transcriptome and Proteome data in between Thalassemia and Control subjects. A) Gene Ontology Molecular Function (GOMF) fold enrichment analysis of Upregulated gene sets in thalassemia. B) Gene set enrichment analysis (GSEA) in the thalassemia group utilizing cluster profiler in R. C) Pathway Enrichment analysis of significant upregulated snRNAs in thalassemia group. D) Gene ontology and molecular function analysis considering the higher abundant proteins in RBC in the thalassemia group than the control. E) STRING analysis showing gene ontology term enrichments for upregulated gene sets. F-G) GSEA analysis of proteome data shows positive normalized enrichment score for: (F) mRNA surveillance and (G) RNA degradation. H) Disease association prediction of low abundant proteins using Metascape analysis tool showing enrichment for RBC and anemia related disorders.

#### snRNA Profile

Transcriptome analysis of red blood cells (RBCs) from healthy individuals and thalassemia patients revealed a notable difference in the distribution of RNA species. Specifically, RBCs from the thalassemia group showed a significantly higher proportion of reads mapping to small nuclear RNAs (snRNAs) compared to the control group (Fig. 1D). This observation suggests an altered RNA composition in the transcriptome of thalassemia RBCs, which may reflect underlying disease-specific regulatory mechanisms. 15 snRNAs that were significantly upregulated in RBCs derived from thalassemia patients. Notably, no significant differences in the expression levels of these snRNAs were observed when comparing the two thalassemia subgroups, indicating that the upregulation of snRNAs is a common feature across the spectrum of thalassemia. To understand the functional relevance of these upregulated snRNAs, a KEGG pathway enrichment analysis was conducted. This analysis revealed that several of the upregulated snRNAs, including RNU5A-1, RNU6-9, RNU6-1, RNVU1-7, RNVU1-18, RNU4-2, RNU1-3, and RNU1-4, were strongly associated with the spliceosome pathway, RNA transport pathway (Fig. 2C). This dual involvement underscores the importance of snRNAs in maintaining RNA processing and transport, processes that may be dysregulated in thalassemia RBCs.

### Proteome Profile

From the mass spectrometry data analysis, a total of 2, 227 proteins were identified, with 826 showing significant differences in abundance (p < 0.05, log2FC > 1 or <-1) between thalassemia patients and controls. Principal component analysis (PCA) and cluster heatmap analysis revealed distinct clustering between the thalassemia and control groups. Notably, DNM1L, RLA2, DNJB4, EIF3B, EIF3D, EIF3H, UBE2H, MIEN1, and EIF3H were the most differentially abundant proteins (Supplementary Table S3 and Fig. 1F-1H). The mass spectrometry proteomics data have been deposited in the ProteomeXchange Consortium via the PRIDE^19^ partner repository with the dataset identifier PXD054385.

Gene Ontology (GO) molecular function fold enrichment analysis of proteomics data revealed that proteins with high abundance in the thalassemia group were significantly enriched in functions related to translation, RNA binding, and unfolded protein binding (Fig. 2D). Gene Set Enrichment Analysis (GSEA) further substantiated these observations by identifying that the upregulated proteins in thalassemia RBCs were prominently involved in the mRNA surveillance pathway and RNA degradation pathways (Fig. 2F-2G and Supplementary Table S3).Conversely, proteins with low abundance in the thalassemia group were enriched in molecular functions such as haptoglobin binding, antioxidant activity, actin binding, and actin filament binding (Supplementary File S1: Fig. S3A). The reduction in these proteins may indicate a compromised ability to manage oxidative stress and cytoskeletal integrity, both of which are crucial for maintaining RBC stability and function. To further explore protein-protein interactions, STRING analysis was performed. This analysis revealed that the highly abundant proteins in thalassemia RBCs were predominantly associated with chaperones and formed intricate interaction networks involving key proteins such as HSPAB, HSPA1B, and DNAJB4 (Fig. 2E). In contrast, the low-abundance proteins were linked to pathways associated with cellular detoxification, particularly those implicated in the pathophysiology of hemolytic anemia (Supplementary File S1: Fig. S3B). This finding aligns with the known oxidative and structural challenges faced by RBCs in thalassemia. Further insights were obtained through Metascape analysis, which revealed that the low-abundance proteins were significantly enriched in pathways and processes related to anemia and RBC disorder-associated diseases (Fig. 2H). This enrichment underscores the relevance of these proteins in maintaining RBC homeostasis and their potential contribution to the clinical manifestations of thalassemia.

#### Transcriptome – Proteome Integrated Findings

To integrate the findings from the RBC transcriptome and proteome, a Venn diagram analysis revealed that 1, 018 protein signatures out of 2, 227 matched with the total transcripts (Fig. 3A). We identified 18 genes that were commonly upregulated and 18 that were commonly downregulated (Fig. 3B). Further investigation into the bioprocess and disease enrichment of these 36 dysregulated genes showed that the most commonly enriched pathways and bioprocesses were involved in translational regulatory machinery, mRNA activation mechanisms, and hydrogen peroxide metabolism processes (Fig. 3C). The expression patterns of the common protein and mRNA signatures were also examined using a heatmap, which showed similar expression trends (Fig. 3D). Consequently, Pearson correlation analysis was performed on the commonly dysregulated genes from the RBC transcriptome and proteome data, revealing that 24 genes exhibited a strong correlation (r > 0.5) between the two omics datasets (Fig. 3E).

**Figure 3.**
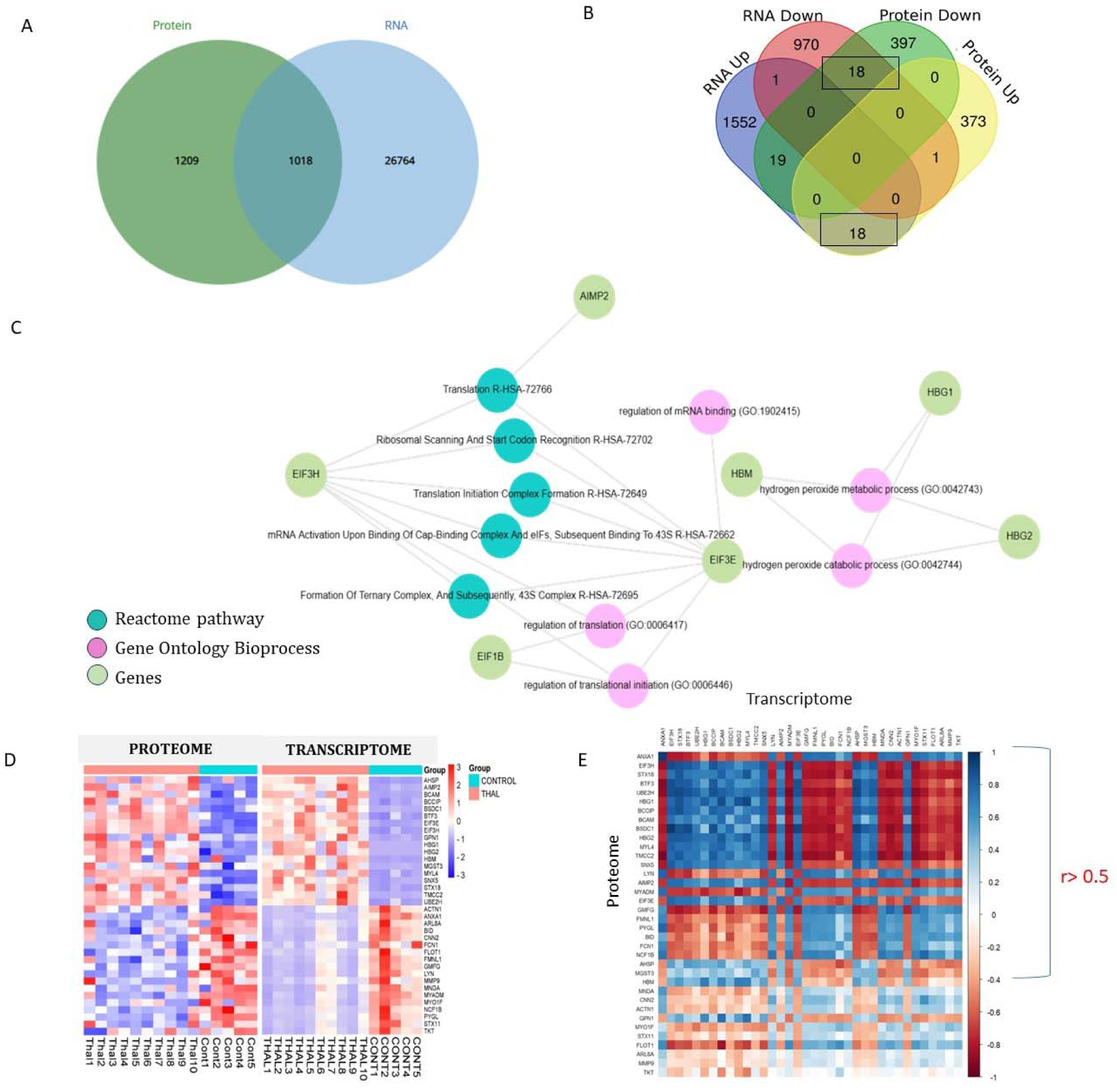
Integration of RBC transcriptome and proteome data: A) Identification of overall matched gene signatures out of 2227 proteins of RBC proteome data. B) Venn Diagram shows common upregulated and down regulated genes from both transcriptome and proteome data. C) GOBP and Pathway enrichment analysis of common altered gene signatures. D) Heatmap showing expression pattern of the common identified genes. E) Correlation of common identified genes of both transcriptome and proteome data.

### Integrated Analysis of Transcriptome and Proteome: Differential Gene Expression Between TDT or severe subjects and NTDT or non-severe Subjects

#### Transcriptome Profile

Comparative transcriptome analysis between the two patient groups—Transfusion-Dependent Thalassemia (TDT) and Non-Transfusion-Dependent Thalassemia (NTDT)—revealed distinct molecular differences. Principal Component Analysis (PCA) and cluster heatmap analysis highlighted significant variations in the transcriptomic profiles of RBCs between these groups (Supplementary File 1: Fig. S2). A total of 579 differentially expressed genes (DEGs) were identified between the TDT and NTDT groups, based on the criteria of an adjusted p-value (FDR) < 0.05 and a log2 fold change (Log2Fc) > 1 or < -1. Among these, C4BPA, CD300H, KRT77, EPB41L3, CLDN5, and AFAR1L2 were the most dysregulated genes, as depicted in the volcano plot (Supplementary File 1: Fig. S2 and Supplementary Table S2). These genes represent key molecular players potentially linked to the differences in disease severity and clinical manifestations between the two thalassemia subtypes.

To uncover factors contributing to the increased severity in TDT patients compared to NTDT, we conducted a detailed analysis of the gene expression profiles. Transcriptome data indicated that upregulated genes in the TDT group were significantly enriched in molecular functions associated with Toll-like receptor binding and RAGE (Receptor for Advanced Glycation Endproducts) receptor binding activity (Fig. 3A). These findings suggest heightened inflammatory and immune response signaling in TDT, which may exacerbate disease severity. In contrast, downregulated genes in the TDT group were enriched in functions related to cytoskeletal protein membrane anchor activity (Supplementary File 1: Fig. S2D). This downregulation could reflect a compromised structural integrity of RBCs, potentially contributing to their reduced lifespan and increased susceptibility to hemolysis in TDT patients.

Gene Set Enrichment Analysis (GSEA) provided additional insights, revealing contrasting pathway activations between the two groups. In the TDT group, genes involved in the apoptosis pathway were significantly upregulated (Fig. 4B), indicating an increased rate of programmed cell death that could exacerbate RBC turnover. Conversely, genes associated with the hematopoietic cell lineage pathway were downregulated (Fig. 4C), suggesting impaired erythropoiesis and a diminished capacity for effective RBC production in TDT. These findings collectively highlight the molecular mechanisms underlying the clinical severity of TDT compared to NTDT. Upregulation of inflammatory signaling and apoptotic pathways, coupled with the downregulation of structural and hematopoietic processes, likely contributes to the greater transfusion dependency and disease burden in TDT patients. These insights provide a foundation for further exploration of targeted therapeutic interventions to mitigate disease severity in thalassemia.

**Figure 4.**
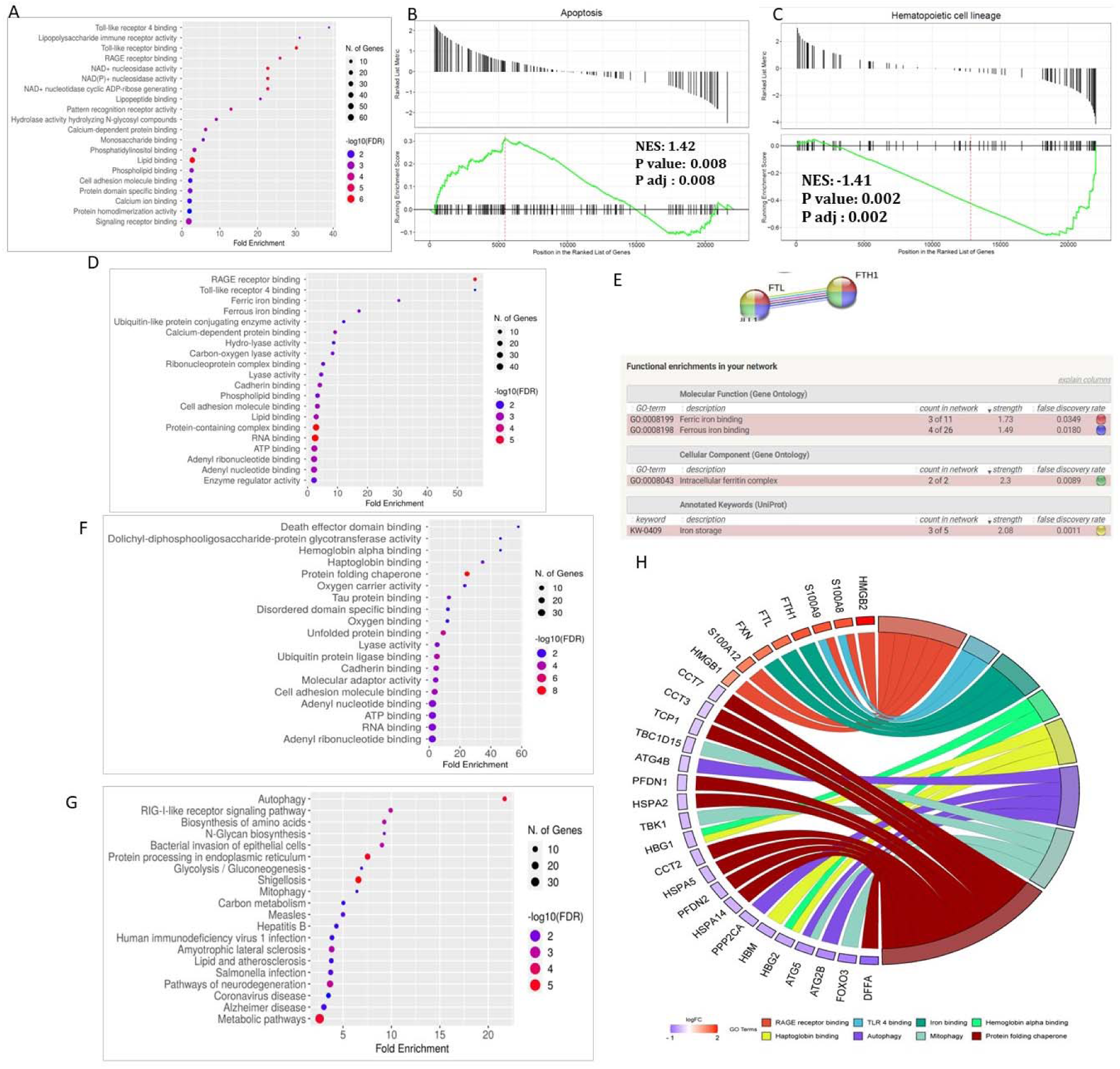
Functional Enrichment Analysis of Transcriptome and Proteome data in between TDT and NTDT subjects. A) GOMF fold enrichment analysis of upregulated gene sets in TDT group. B) & C) Gene set enrichment analysis (GSEA) for DEGs between TDT and NTDT groups using cluster profiler in R. Genes associated with (B) apoptosis were upregulated and (C)hematopoietic cell lineage were downregulated. D) GOMF fold enrichment analysis of high abundant proteins in TDT. (E) STRING analysis for upregulated gene sets in TDT. (F) GOMF fold enrichment analysis of low abundant proteins in TDT. (G) KEGG pathway fold enrichment analysis for low abundant protein sets in TDT. (H) GO: Circos chord plot showing high and low abundant proteins and their enriched molecular functions and pathways that might be associated with disease severity.

#### Proteome Profile

PCA analysis revealed a 43% variance at PC1 between the TDT and NTDT groups. A total of 430 differentially abundant proteins were identified between the two patient groups (Supplementary Table S4). The volcano plot indicated that HMGB2, LAT4, LKHA4, MP2K2, DYL1, EXOC1, RO52, and ACL6A were the most differentially abundant proteins between the TDT and NTDT subgroups (Supplementary File 1: Fig. S4). For the proteome data, a log fold change threshold of >0.5 or <-0.5 at a p-value < 0.05 was established. The proteome data analysis showed that highly abundant proteins in TDT were enriched for Toll-like receptor and RAGE receptor binding activity (Fig. 4D). Conversely, low-abundance proteins were enriched for hemoglobin alpha binding, haptoglobin binding, oxygen carrier activity, protein folding chaperone activity, and oxygen binding (Fig. 4F). KEGG pathway analysis indicated that low-abundance proteins were involved in pathways related to autophagy and mitophagy (Fig. 4G). Furthermore, STRING network analysis revealed that FTH1 and FTL proteins were found in higher abundance in TDT compared to NTDT, indicating their association with the ferroptosis pathway. This analysis also demonstrated significant iron storage in TDT (Fig. 4E). The Gene Ontology circos chord plot illustrated the altered proteins identified in transfusion-dependent thalassemia (TDT) and their connection with molecular functions and pathways potentially contributing to the increased severity in TDT patients(Fig. 4H).

The Gene Ontology circos chord plot highlighted the altered proteins identified in transfusion-dependent thalassemia (TDT) and their associations with molecular functions and pathways contributing to the increased severity observed in TDT patients. The analysis revealed critical roles of RAGE receptor binding, TLR4 binding, iron binding, hemoglobin alpha binding, haptoglobin binding, autophagy, mitophagy, and chaperone-mediated protein folding in the pathophysiology of thalassemia. These findings were derived from gene ontology enrichment analysis and STRING protein-protein interaction network analysis.

Specifically, upregulated proteins in TDT included S100A8 and S100A9, which were linked to TLR4 binding, while HMGB2, S100A8, S100A9, S100A12, and HMGB1 were associated with RAGE receptor binding. Proteins such as FTH1, FTL, and FXN were enriched for iron-binding functions. Conversely, downregulated proteins in TDT compared to NTDT were associated with several critical pathways. HBG1 and HBG2 were linked to hemoglobin alpha binding, while HBG1, HBG2, and HBM were connected to haptoglobin binding. Proteins such as ATG4B, PPP2CA, ATG5, and ATG2B were involved in autophagy, while TBC1D15, TBK1, ATG5, and FOXO3 were associated with mitophagy. Furthermore, proteins like CCT7, CCT3, TCP1, PFDN1, HSPA2, CCT2, HSPA5, PFDN2, HSPA14, and DFF4 were implicated in chaperone-mediated protein folding (Fig. 4H).

A statistical integration of the transcriptome and proteome data of dysregulated genes between TDT and NTDT was conducted. According to the Venn diagram, 10 genes were commonly altered between the TDT and NTDT groups (Supplementary File 1: Fig. S5A). Furthermore, Pearson correlation analysis of these genes showed a high correlation coefficient.

### Identifying Altered Expression of snRNPs in RBC

Among the RBC proteome data, 72 proteins were identified as snRNPs according to the UniProt database. Of these, 22 snRNPs showed altered expression (log2 fold change >1 or <-1, p < 0.05), with 19 upregulated in thalassemia patients (Supplementary Table S5). These snRNPs, which include small nuclear ribonucleoproteins and U1-U6 snRNA-associated Sm-like proteins, were highly enriched for post-transcriptional modification regulation and small nuclear RNP binding. KEGG pathway analysis highlighted enrichment in spliceosome and RNA degradation pathways (Fig. 5A-5D).

**Figure 5.**
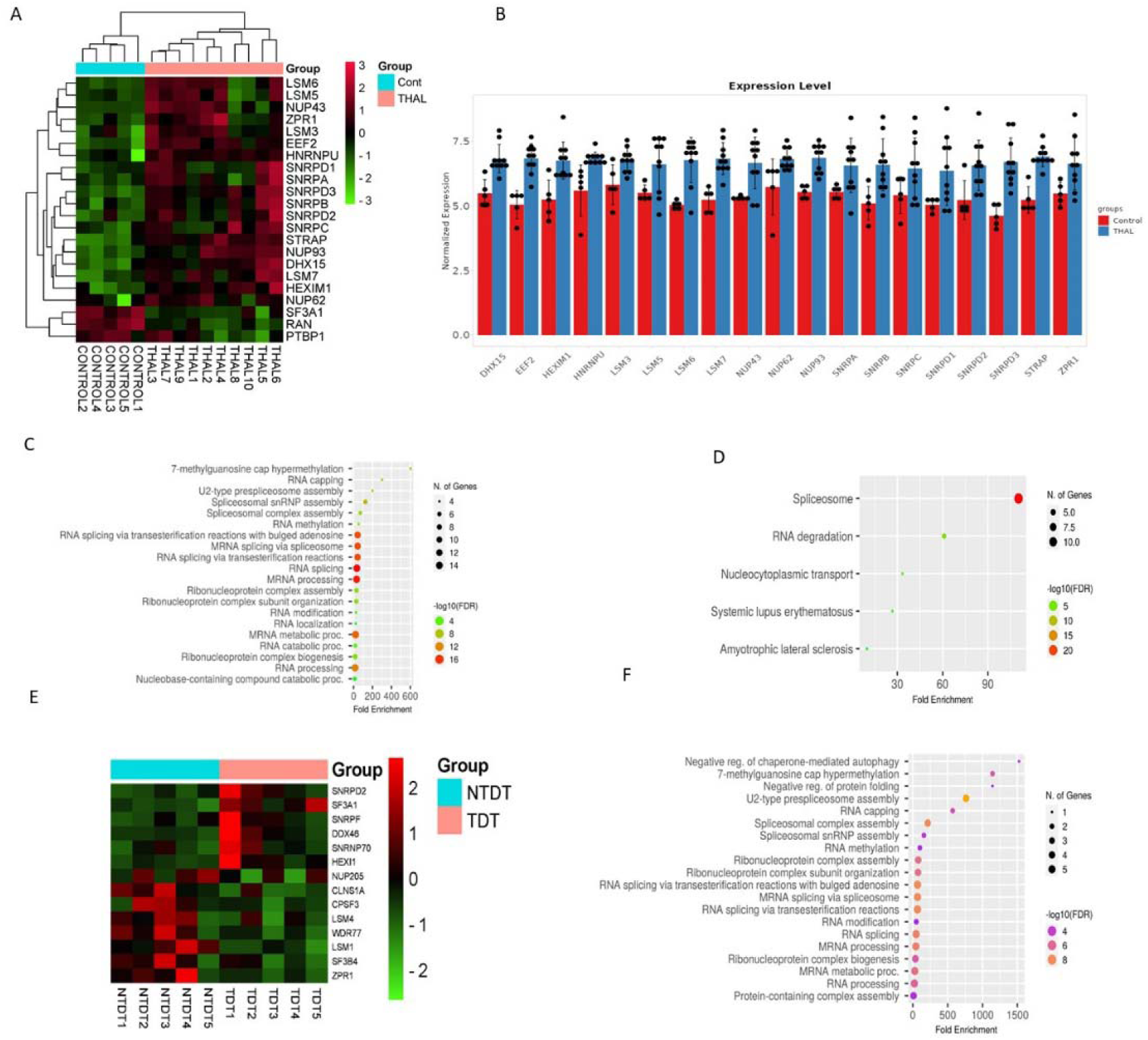
Analysis of altered expression signatures of snRNPs in RBC proteome data: A) Heatmap showing snRNPs were more highly expressed in thalassemia subjects compared to control subjects. B) Bar graph representing normalized expression levels of snRNPs in thalassemia and control groups. (C) & (D) Fold enrichment analysis of upregulated snRNPs in thalassemia: (C) GOBP enrichment and (D) KEGG pathway enrichment. E) Heatmap showing altered expression of snRNPs between TDT and NTDT groups. F) GOBP fold enrichment of upregulated snRNPs in TDT.

Additionally, 14 spliceosome or mRNA processing-associated proteins showed altered expression in TDT patients compared to NTDT patients, with 6 snRNPs upregulated in TDT (p-value <0.05, log2Fc >0.5 or <-0.5). Interestingly, these upregulated snRNPs were enriched for the negative regulation of autophagy, methylation processes, and protein folding. Conversely, 8 snRNPs were downregulated in TDT, implicating them in splicing processes, splice site selection, polyadenylation, and mRNA decay (Fig. 5E-5F).

## DISCUSSION

In this study, we analysed the red blood cell (RBC) molecular profiles of thalassemia patients with the β0/β^+^ genotype of the HBB gene. Despite sharing the same genetic background, we selected two clinical distinct groups of patients. The first group consists of those with a severe phenotype, also known as transfusion-dependent thalassemia (TDT), who require regular blood transfusions due to consistently low baseline hemoglobin levels. These frequent transfusions lead to clinical issues like iron overload, so most of these patients are undergoing iron chelation therapy to manage excess iron.

In contrast, the second group includes patients with the same genetic mutations but a milder, non-severe phenotype, referred to as non-transfusion-dependent thalassemia (NTDT). These patients do not need regular blood transfusions, and most do not suffer from iron overload or require chelation therapy. For this group, clinical management focuses on monitoring their condition, ensuring proper nutrition, and avoiding factors that could worsen anemia.

To find out the influence of non-genomic factors, known genomic modifiers—such as deletions in the alpha-globin chain or the γG-globin -185 (C>T) polymorphism, which can affect disease severity—we ensured that none of the enrolled patients had these modifiers.

In the initial step of this present investigation we explored both the transcriptome and proteome profiles of RBC from Thalassemia patients to understand the underlying causes of the phenotype profiles for Thalassemia condition than normal state. Subsequently, we compare between severe and non severe group of Thalassemia. We first analysed the transcriptome data, and to confirm or validate the findings of transcriptome, we further investigated the proteome from the RBC of the same patients.

We first analysed the transcriptome data. The upregulated genes in thalassemia compared to normal volunteers showed activation of different globin gene activities including hemoglobin binding, oxygen carrier activity and hemoglobin alpha binding, Further, Gene Set Enrichment Analysis (GSEA), indicated the enrichment of protein folding chaperones in Thalassemia. KEGG enrichment analysis revealed that the upregulated DEGs were associated with ferroptosis. The downregulated gene sets in RBCs of thalassemia were significantly enriched in IgG binding, interleukin-1 receptor binding, Pattern recognition receptor binding, Phosphotyrosine residue binding, Phosphoprotein binding, Actin filament binding and Actin binding.

On investigating the proteome data from Thalassemia and control subjects, we found that most of the transcriptional findings were validated. STRING analysis revealed that in thalassemia, highly abundant proteins are associated with chaperone activity. Based on gene set enrichment analysis (GSEA) we discovered that upregulated proteins are also associated with mRNA surveillance and RNA degradation pathways in thalassemia. Conversely, low abundant proteins were highly enriched in anemia and RBC disorder related diseases in thalassemia, along with haptoglobin binding, antioxidant activity, and actin binding activities.

Examining the overall similarities between the transcriptome and proteome data, Gene Ontology molecular function, GSEA, and STRING analyses revealed that upregulated gene sets and highly abundant proteins were predominantly associated with protein folding chaperone activity in Thalassemia patients compared to controls (Fig. 4). Notably, Hsp70 and Hsp90 family genes, which mediate stress-induced HRI activation in erythroid cells and are associated with globin gene translation, showed increased expression ^20–22^. Specifically, HSPA8 and HSPAB1 genes were significantly more abundant at the translational level in Thalassemia patients. Conversely, gene sets and proteins linked to actin filament binding activity were downregulated (Supplementary File S1: Fig. S1A and S3A). Actin filaments are crucial for red blood cell maturation and membrane behavior, which inhibit redox action in mature RBCs ^23^. It has also been reported that ACTIN1 and LYN are associated with the terminal erythropoiesis stage ^24–26^. These findings suggest that disruptions in these genes contributes to Thalassemia pathogenesis by affecting erythroid homeostasis as well as redox action. Therefore, the most significantly altered genes in the RBCs of thalassemia patients are involved in RBC structure, protein folding, or apoptosis which are responsible for the pathological severity.

In this multi-omics investigation, we selected thalassemia patients with the same HBB genotype, specifically compound heterozygous for IVS-1-5 (G>C) and CD 26 (G>A), who also lack mutations in the alpha globin gene. This choice ensures that the severity of the condition is not influenced by the mutational burden, allowing us to focus on other molecular factors. Consequently, these cohorts exhibit differences in disease severity that are not due to globin gene mutation load but rather other molecular influences. We hypothesize that non-severe or NTDT patients may possess comparative better molecular pathways that protect them from more severe ineffective erythropoiesis. A comparative transcriptome and subsequent proteome analysis between the two groups could provide insights into potential drug targets.

We analysed and compared the differential expression between the TDT and NTDT groups. Reanalysis of the transcriptome data indicated that some dysregulated genes in TDT are associated with apoptosis, while downregulated genes are linked to the hematopoietic cell lineage. These results correlate with the higher incidence of ineffective erythropoiesis in TDT subjects (Fig 4B & 4C). STRING interaction analysis of the proteome data identified FTH1 and FTL as highly abundant proteins in TDT, indicating higher iron deposition and lower oxygen carrier activity in these patients (Fig. 4E). Elevated iron levels can lead to ferroptosis and organ damage, suggesting ferroptosis as a potential mechanism contributing to TDT severity ^27–28^.

In TDT, low-abundance proteins are enriched for globin binding, protein folding chaperones, oxygen carrier activity, antioxidant activity, autophagy, and mitophagy pathways (Fig. 4F and 4G). In our protein profile, HBG was found to be significantly reduced in TDT patients compared to NTDT, alongside elevated level of PIP4K2A. According to previous reports, PIP4K2A catalysis the breakdown of phosphatidyl inositol 5 phosphate (PI5P), which, in turn, helps increase HBG gene expression ^29–30^. Therefore, the increased protein levels of PIP4K2A might lead to the enhanced breakdown of PI5P. Autophagy and mitophagy-related proteins, including FOXO3, ATG2B, ATG4B, and ATG5, were consistently reduced in most TDT patients. Additionally, reduced levels of protein-folding chaperones such as HSPA5, PFDN1, TCP1, HSPA2, CCT7, PFDN2, DFFA, CCT3, CCT2, and HSPA14 were observed in TDT patients with the same genotype as NTDT patients (Fig. 4H). Both processes crucial during late-stage erythropoiesis. Hence, the dysregulation of these functional proteins might contribute to the severity of the phenotype and the increased demand for blood transfusion.

It has also been found in both transcriptome and proteome data analysis that toll-like receptor (TLR) activity and RAGE receptor activity are upregulated in TDT patients. Due to multiple blood transfusions, TDT patients are at a higher risk of encountering foreign antigens that activate the TLR pathway, initiating an innate immune response. Recent studies have emphasized bacterial infections in hemolytic diseases ^31^. Significant quantities of damage-associated molecular patterns (DAMPs) released during hemolysis can activate various inflammatory and TLR signalling pathways ^32^. RAGE receptor activity is also associated with the inflammatory response ^33^. Overall, this evidence suggests that TDT patients experience increased inflammation, worsening the disease condition (Fig. 4A & 4D)

Interestingly, we have found a global upregulation of snRNAs in Thalassemia. Most of these snRNAs are members of the RNA spliceosome complex. There were an increased percentage of snRNA reads across all patient groups, identifying 15 snRNAs with elevated expression. Several reports have shown that altered expression of snRNAs is associated with transcriptional inhibition and post-transcriptional alterations^34^. Accordingly, snRNA RNU4-2, involved in tri-snRNP complex assembly along with U6 snRNA, a core component of the spliceosome, is essential for proper splicing. Dysregulation of these RNAs can disrupt splicing during erythropoiesis ^35^. RNVU1-7, part of U1 snRNP, binds pre-mRNA 5’-splice sites, and its dysregulation can lead to splicing defects, impacting cell maturation ^36^. RNU6ATAC, part of the minor spliceosome, is linked to ineffective erythropoiesis ^37–38^. According to earlier reports, upregulation of RN7SK can hinder the release of RNA polymeraseII and is associated with thalassemia pathogenesis ^39–40^. GSEA analysis of differentially abundant proteins in thalassemia showed enrichment of the mRNA surveillance and RNA degradation pathway (Fig 2F-2G). The proteomics result showed that 19 snRNPs were upregulated in thalassemia compared to controls and 14 snRNPs were dysregulated in TDT compared to NTDT. Most of these snRNPs are involved in post transcriptional modification, spliceosome formation and the mRNA degradation pathway. We noticed HEXIM1, one of the snRNPs, was upregulated in the TDT group. HEXIM1, part of the 7SK complex, regulates the pausing of RNA polymerase II. Pausing of polymerase II interferes with terminal erythroid maturation ^41–42^. Thus, the HEXIM1-RN7SK complex components in TDT patients augment ineffective erythropoiesis and are also responsible for the increased hemolytic anemia in TDT patients.

It has been observed that SF3B4, WDR77, and ZPR1, along with other snRNPs, are downregulated in TDT patients (Supplementary Table S5). SF3B4 is a member of the SF3B family of splicing factors and constitutes part of U2snRNP. Previous studies have shown that blocking SF3B leads to the accumulation of unspliced pre-mRNAs^43^. Therefore, the downregulation of SF3B4 in TDT, as observed in our study, may significantly contribute to altered splicing or splicing inhibition of the beta-globin gene or other crucial erythroid genes, potentially resulting in a more severe phenotype. In previous studies, it has been demonstrated that the loss of WDR77 results in alternative splicing of numerous crucial genes, along with an increase in the expression of apoptotic genes. RNA processing genes were notably impacted by the absence of WDR77. Our study revealed a downregulation of WDR77 in TDT. This downregulation potentially leads to aberrant splicing. Furthermore, the upregulation of apoptotic genes such as CASP6 and BID in our proteome data indicates an increase in apoptosis^44^. ZPR1 is a bespoke chaperone involved in the biogenesis of eEF1A1 factor, in its depletion, ZPR1 results in misfolded eEF1A1. Misfolded eEF1A1 dysregulates HSP 70 transcription, translation and stabilization ^45^, potentially diminishing chaperone mediated autophagy in TDT. Conversely, we observed upregulation of snRNP70 in TDT. It has been reported that upregulation of snRNP70 can reduce chaperone mediated autophagy ^46^. Moreover, the dysregulation of snRNPs in TDT patients are responsible for the negative regulation of chaperone mediated autophagy, and the protein folding process which are enhanced in TDT patients (Fig 4D). Chaperones play protective roles in preventing apoptosis, facilitating hemoglobin synthesis in erythroid precursors, and maintaining hematopoietic stem cell function ^47–48^. Therefore, it is obvious that the reduced expression of autophagy-related functional proteins, chaperones and the snRNPs mediated negative regulation of autophagy and protein folding process appear to contribute to the more severe phenotype in TDT patients, who require regular transfusions, compared to NTDT patients with the same genotype.

Based on our comprehensive analysis, we identified key protein signatures and their associated pathways that contribute to disease severity in TDT, as illustrated in Figure 6. The critical protein signatures and their target pathways are as follows:

1. Increased PIP4K2A Proteins: This leads to a reduction in gamma globin levels.
2. Altered Expression of snRNA/snRNP Complex HEXIM1 and Other snRNPs (SNRNP70, SF3B4, WDR77): These changes impact transcription, translation, and aberrant splicing
3. Reduced Levels of Functional Autophagy Regulatory Proteins (ATG2B, ATG4B, ATG5, FOXO3).
4. Decreased Abundance of Functional Hsp70 Heat Shock Chaperone Proteins (HSPA2, HSPA14).
5. Increased Levels of Iron Regulatory Proteins (FTH1, FTL) in TDT Patients.

**Fig 6.**
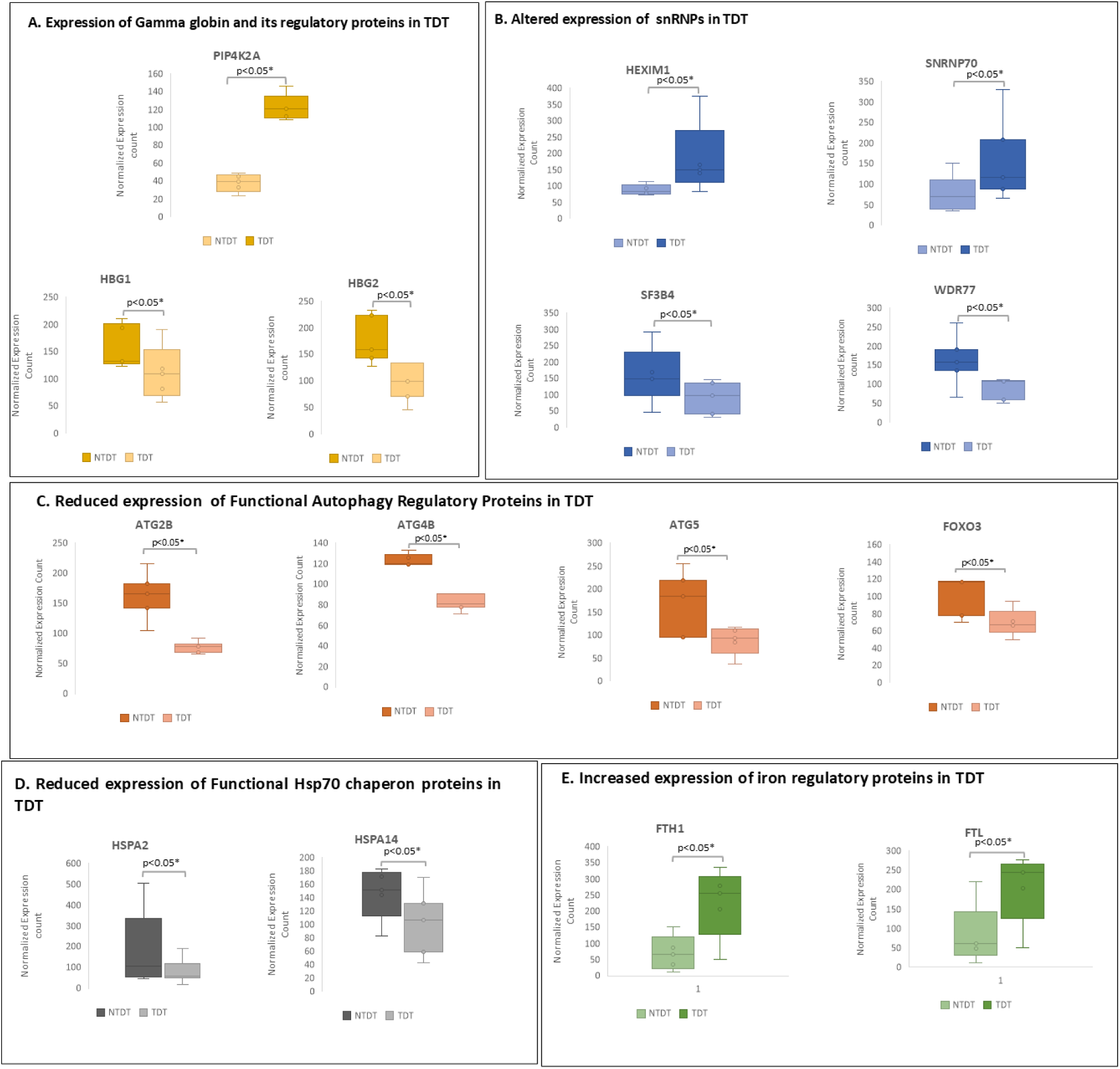
Differential abundance of different protein between TDT and NTDT group linked with key pathways: A) Gamma globin production: Increased expression of PIP4K2A proteins and reduction of HBG1 and HBG2 in TDT group, B) Transcription regulation and splicing: Higher abundance of HEXIM1 SNRNP70 and down regulation of, SF3B4, WDR77 in TDT group C) Autophagy regulatory proteins: Higher abundance of ATG2B, ATG4B, ATG5, FOXO3 in TDT, D)Chaperon and protein folding: Reduced abundance of functional Hsp70 heat shock chaperon proteins: HSPA2, HSPA14. E) Iron Regulatory pathway: Higher abundance of FTH1, FTL in TDT patients.

These findings highlight the intricate molecular mechanisms causing higher percentage of ineffective erythropoiesis or anemia, iron overload linked with clinical severity.

To the best of our knowledge, this is the first study to analysed the potential drug target for thalassemia with comprehensive analysis of RBC directly taking from patients. In this study, we investigated the transcriptome and validated it through the proteome of RBCs from an equal number of non-severe and severe thalassemia patients to address the causes of disease severity. Our goal was to identify molecular targets beyond the mutations in the beta-globin gene. We focused on RBCs, as they are the most affected cellular system in thalassemia. Our results indicate that the traditionally believed lower expression of the gamma-globin gene (HbF) is not the sole contributor to severity. Although targeting HbF induction is a major focus in thalassemia drug development. Over all through these study, we have identified first time non genomic targets that can be exploited for drug development (Figure 7): 1) Increased expression of PIP4K2A, which decrease PI5P, and triggers the lowering of Gamma-1-globin or Gamma-2-globin (HBG) in TDT compared to NTDT. Targeting PIP4K2A might be the new drug target: 2) Severe group exhibit upregulation of snRNA/snRNP complex: HEXIM1-RN7SK. This complex can hold the RNA Pol II, which can lead to aberrant or reduced transcription which may induce truncated protein and or reduced translation and increased ineffective erythropoiesis. HEXIM1-RN7SK can be the target of choice, 3) Downregulation of SF3B4 and WDR77 in TDT associated with aberrant splicing and that can lead to reduced protein synthesis or protein misfolding respectively. Thus, activation of WDR77 can be another route of drug target. 4-5) Reduced expression of chaperone family proteins further leading to higher state of protein misfolding and generate ROS. Decreased expression of autophagy linked protein in TDT can lower the autophagy, which again increase ROS and ultimately increase haemolysis. Thus autophagy inducer can be used for thalassemia treatment. 6) Increase hemolysis can induce higher expression iron binding protein: FTH1, FTL can also leads to ferroptosis and maturation arrest and augment ineffective erythropoiesis in TDT, Reduced autophagy/mitophagy along with upregulation of proapoptotic proteins can increase RBC destruction. Thus FTH1, FTL can be drug target for thalassemia treatment ^49^.

**Figure 7.**
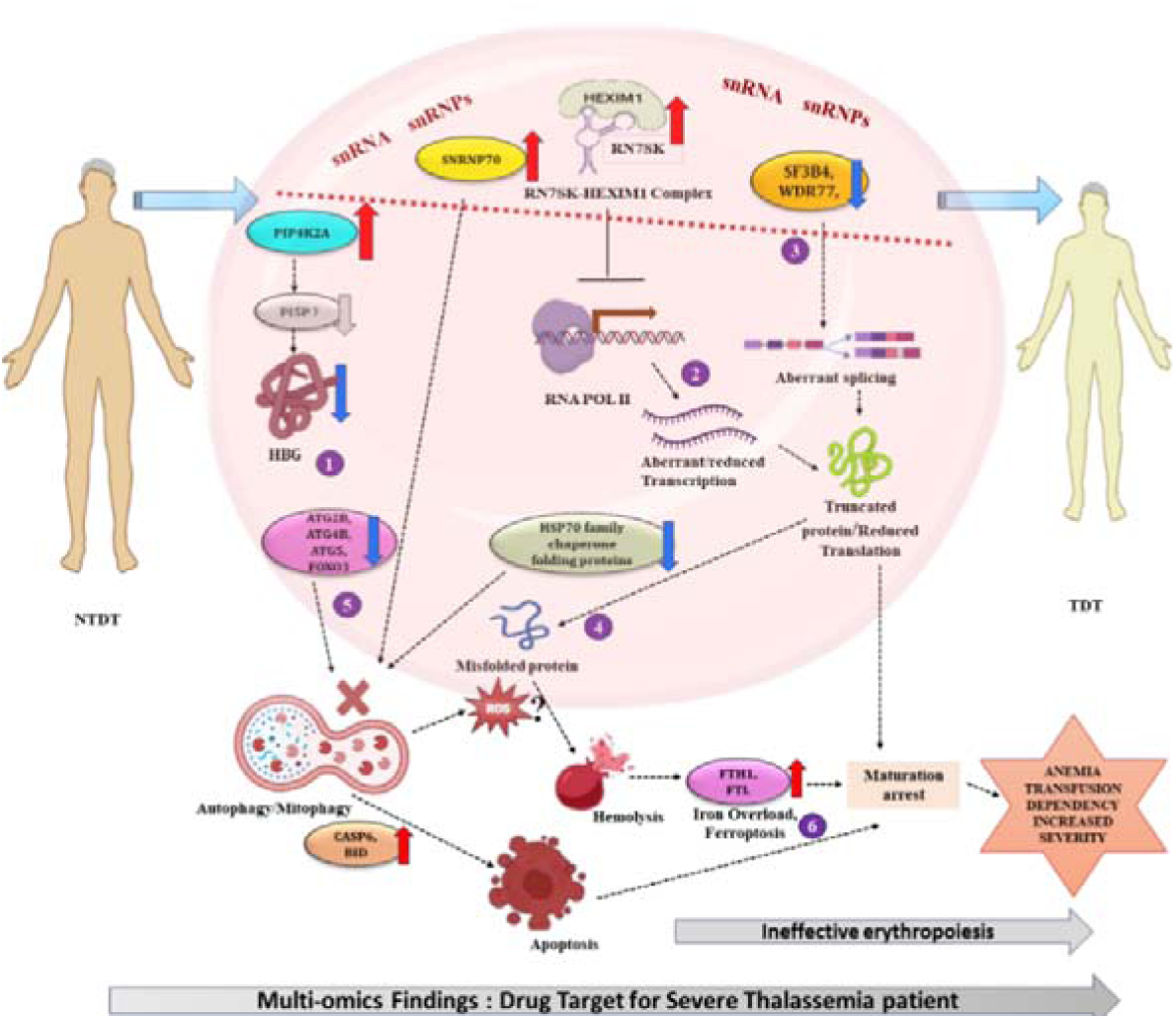
Graphics showing different molecular events comprising snRNA-snRNP complex along with different molecular pathway that happen inside the RBC which are responsible for thalassemia severity linked with augmented ineffective erythropoiesis and anemia as potential drug target. 1) Increased expression of PIP4K2A, which decrease PI5P and triggers the lowering of Gamma-1-globin or Gamma-2-globin (HBG) in TDT compared to NTDT: 2) Severe group exhibit upregulation of snRNA/snRNP complex: HEXIM1-RN7SK. This complex can hold the RNA Pol II, which can lead to aberrant or reduced transcription which may induce truncated protein and or reduced translation and increased ineffective erythropoiesis., 3) Downregulation of SF3B4 and WDR77 in TDT associated with aberrant splicing and that can leads to reduced protein synthesis or protein misfolding respectively. 4-5) Reduced expression of chaperone family proteins further leading to higher state of protein misfolding and generate ROS (4). Decreased expression of autophagy linked protein in TDT can lower the autophagy, which again increase ROS and ultimately increase hemolysis (5). 6) Increase hemolysis can induce higher expression iron binding protein: FTH1, FTL can also leads to ferroptosis and maturation arrest and augment ineffective erythropoiesis in TDT, Reduced autophagy/mitophagy along with upregulation of proapoptotic proteins can increase RBC destruction in TDT.

The RBC is the most potent system used for Thalassemia drug development^8^. Thus, molecular dysregulated pathways discovered in this present study, as directly captured from the RBC can be further been targeted individually with the different inhibitors those are under trials of the respective dysregulated pathways ^50–54^. These findings may also promote future research or trials for thalassemia treatment by the repurposing of the existing drugs already used for the treatment of other diseases. Overall, the key impact of the present research could lead to the discovery of new treatments for hemoglobinopathies or thalassemia. Finally, this study serves as a pioneer effort to unravel the molecular intricacies influencing phenotypic variations in monogenic disorders beyond the impact of mutational burden.

## MATERIALS & METHODS

### Cohort Design and Sample Collection

A cohort of 285 thalassemia patients, aged 8 to 20, with the β0/β+ genotype, was screened based on stringent criteria, focusing on individuals with the compound heterozygous HBB beta-globin genotype IVS 1-5 (G>C) and CD 26 (G>A). All participants had intact spleens. From this group, ten individuals were selected. According to the Thalassemia International Federation (TIF) guidelines on baseline hemoglobin levels and transfusion requirements, five patients were classified as transfusion-dependent thalassemia (TDT), or severe thalassemia, requiring transfusions every two to three months^16^. The remaining five were categorized as non-transfusion-dependent thalassemia (NTDT), or non-severe thalassemia, not requiring frequent transfusions. Clinical details for each patient are provided in Supplementary Table S1. Additionally, five healthy donors in the same age group were included as controls. Peripheral blood (4.5-5 mL) was collected in EDTA vials from each participant. The experimental workflow has been presented in Visual abstract. The study received approval from the Institutional Clinical Ethics Committee of The University of Burdwan. The study was carried out following the WMA Declaration of Helsinki and adhering to ethical principles for medical research involving human subjects. As part of a larger multicentric project, written consent was obtained from all enrolled subjects. The study did not interfere with patient care and treatment.

### RBC Isolation and RNA Extraction

Red Blood Cells (RBCs) were isolated from peripheral blood using density gradient separation. RNA was extracted using the TRIZOL Plus RNA purification kit (Invitrogen Cat No. 12183555) and assessed for quality and quantity.

### RNA Sequencing and data analysis

RNA sequencing libraries were prepared using the KAPA RNA Hyper Prep kit (Kapa Biosystems, Cat No. KK8561) and sequenced on the NOVA SEQ 6000 Illumina Platform utilizing 2X100bp read length targeting 75 million reads per sample at the NIBMG genomics facility. Total RNA sequencing were carried with 5 Biological replicates from each group.

Raw sequencing data underwent quality control checks. Paired-end reads were joined and mapped to the hg 38 reference genome using Illumina DRAGEN V.4. BAM files were used to generate expression read counts. Differential Gene Expression (DGE) analysis was performed with DESEQ2 (R, Bioconductor package) to produce fold change in gene expression [LOG**_2_**] between the groups. To determine significant changes in gene expression log**_2_**FC >1 or < -1 and adjusted P Value < 0.05 were set as cutoff limits. The BioMart (Ensemble) web application was used to annotate the non-Coding RNA of the transcript.

### Sample Preparation for RBC Proteomics Study

A 75µL suspension of RBC cells was used to prepare RBC lysate. Proteins were extracted with RIPA buffer (Thermo Fisher Scientific), and hemoglobin was depleted using the HemoVoid kit (BSG, Cat No. HVB-MS10). Protein concentration was measured with the BCA assay kit (Thermo Fisher Scientific). 200 μg of protein was processed with 50-mM ammonium bicarbonate buffer, reduced with DTT, alkylated with iodoacetamide, and hydrolyzed with MS Grade trypsin at 37°C for 18 hours.

### LC MS/MS Analysis

The LC MS/MS platform was set up as described in a previous study ^17^. LC MS/MS was performed using an RSLC nano system coupled to a QExactive Plus Orbitrap MS. A 750 ng digest was separated on a C18 analytical column (2 μm, 100 Å particles, 75 μm × 50 cm) with a gradient separation using solvent system A (0.1% formic acid (FA), 3% acetonitrile in water) and solvent B (0.1% FA in 80% acetonitrile). The data was acquired in positive-ion mode and analysed by QExactive in dd-MS2 mode, with one full MS scan (resolution 140000 at m/z 200). The acquired RBC proteomic datasets were processed by SEQUEST HT in Proteome Discoverer 2.4 (Thermo Fisher Scientific). The search parameters included variable modification of methionine oxidation and fixed modification of cysteine carbamidomethylation. Peptide identification was performed using a 10-ppm precursor ion tolerance; the product ion tolerance was 0.05 Da. Peptide spectrum matches were adjusted to 1% FDR. Identified peptides were annotated to the protein with a UniProt human database. snRNPs were manually filtered using the Uniprot snRNP database from the proteomics data. Raw abundance values were analysed using the DEQMS package for protein differential abundance between normal vs. thalassemia and severe (TDT) vs. non-severe (NTDT) thalassemia.

### Statistics and Bioinformatics Analysis

Statistical analysis was conducted using GraphPad Prism V.10 for both transcriptome and proteome data. For sample size power estimation online RNA seq power calculation tool was utilized. Student t-test (p <0.05) was used for box plot analysis of Key significant proteins identified in between TDT and NTDT group. Gene Set Enrichment Analysis (GSEA) was performed using the clusterProfiler 3.16 package (Bioconductor R) to identify the most enriched up- or down-regulated functional pathways. Gene Ontology, protein-protein interaction, and disease ontology fold enrichment analysis were conducted using ShinyGO, EnrichR, STRING, and Metscape 2.0 ^18^ webtools, respectively.

## ETHICAL APPROVAL

The study received approval from the Institutional Clinical Ethics Committee of The University of Burdwan (Approval No. IEC/BU/2021/10).

## DATA AVAILABILITY

All transcriptome data are submitted to NCBI -SRA database with Bio project accession no. PRJNA1074078. Proteome data are submitted to PRIDE archive database; Project accession no: PXD054385, all these data will be publicly available as per procedure.

## Data Availability

All transcriptome data are submitted to NCBI -SRA database with Bioproject accession no. PRJNA1074078. Proteome data are submitted to PRIDE archive database; Project accession no: PXD054385, All data produced in the present study are available upon reasonable request to the authors

## ACKNOWLEDGEMENTS

All the thalassemia patients and control subjects. National Institute of Biomedical Genomics, Kalyani, for National Genomics core (NGC) for RNAseq and Proteomics Facility for proteomics work, and The University of Burdwan, West Bengal for necessary support. The present work was partially supported by funding of Department of Biotechnology, Govt. of India [Sanc. No-BT/PR26461/MED/12/821/2018], intramural grant support of National Institute of Biomedical Genomics (NIBMG), Kalyani.

